# Dragon III- Phase II Randomized Controlled Trial: Neoadjuvant FLOT versus SOX for Patients with Locally Advanced Gastric Cancer

**DOI:** 10.1101/2020.06.21.20136887

**Authors:** Birendra Kumar Sah, Benyan Zhang, Huan Zhang, Jian Li, Fei Yuan, Tao Ma, Min Shi, Wei Xu, Zhenglun Zhu, Wentao Liu, Chao Yan, Chen Li, Bingya Liu, Min Yan, Zhenggang Zhu

## Abstract

**Background:** Neoadjuvant chemotherapy with docetaxel, oxaliplatin, fluorouracil, and leucovorin (the FLOT regimen) has shown promising results in terms of pathological response and survival rate. However, tegafur gimeracil oteracil potassium capsule (S-1) plus oxaliplatin (the SOX regimen) is a more favorable chemotherapy regimen in Eastern countries. We conducted this study to evaluate the safety and efficacy of both regimens and to explore a suitable regimen for gastric cancer patients.

**Methods:** Patients with locally advanced gastric cancer (LAGC) were 1:1 randomly assigned to receive either 4 cycles of the FLOT regimen or 3 cycles of the SOX regimen before curative gastrectomy. The primary endpoint was the comparison of complete or subtotal tumor regression grading (TRG1a+ TRG1b) in the primary tumor.

**Results:** Altogether, 74 patients were enrolled between August 2018 and March 2020. There was no significant difference in pretreatment clinicopathological parameters between the FLOT group and the SOX group (p>0.05). There was no significant difference in adverse effects or postoperative morbidity and mortality between the two groups (p>0.05). Similarly, there was no significant difference in the proportion of tumor regression grading between the FLOT group and the SOX group (p>0.05). In the ITT population, complete or subtotal TRG was 20.0% in the FLOT group versus 32.4% in the SOX group (p>0.05).

**Conclusion:** Our study demonstrates that the FLOT and SOX regimens are similarly effective for locally advanced gastric cancer patients in terms of clinical downstaging and pathological response. A large-scale phase III randomized controlled trial is necessary to validate this result.

## Background

For locally advanced gastric cancer (LAGC), there has been a positive trend in neoadjuvant chemotherapy after the milestone publication of the MAGIC trial in 2006, and results from this trial were recently even supported by those from clinical trials from Asian countries (1-5). Furthermore, recent studies have shown that neoadjuvant chemotherapy is well tolerated and does not influence postoperative morbidity or mortality in gastric cancer patients (6). A large-scale German study clearly showed the superiority of neoadjuvant docetaxel, oxaliplatin, fluorouracil, and leucovorin (the FLOT regimen) over epirubicin, cisplatin, and fluorouracil or capecitabine (the ECF or ECX regimens, respectively) in terms of pathological response and overall survival (7, 8). The FLOT regimen is not a common chemotherapy in China; however, there have been published studies that show that the modified or standard FLOT regimen is safe and effective in Chinese patients (9, 10). Taxane-based triplet chemotherapy was considered more toxic in the past; therefore, doublet chemotherapy with the oral tegafur-gimeracil-oteracil potassium capsule (S-1) is the mainstream adjuvant chemotherapy in Asian countries, and a few studies have suggested S-1 plus platinum-based chemotherapy as neoadjuvant chemotherapy for locally advanced gastric cancer, especially with bulky lymph nodes(11-13).

In recent years, several studies have been carried out in East Asian countries on the efficacy of perioperative chemotherapy for patients with LAGC. Among them, the preliminary results of two large-scale RCT trials (RESOLVE, RESONANCE) in China suggested that the neoadjuvant SOX regimen is beneficial in terms of R0 resectability, TRG, ypTNM, and pCR. Patients in the neoadjuvant chemotherapy group achieved a longer 3-year DFS than the control group (14, 15). To our knowledge, there is no previous study on neoadjuvant chemotherapy for LAGC that compared the efficacy between the SOX and FLOT regimens. For patients undergoing neoadjuvant chemotherapy, the pathological response rate or tumor regression grading is considered one of the major factors that influence overall survival (16, 17). Therefore, the main purpose of this study was to compare the rate of postoperative tumor regression between the neoadjuvant chemotherapy FLOT and SOX groups.

## Methods

This was an investigator-initiated, phase II, open-label, randomized controlled trial. All patients were enrolled between August 2018 and March 2020 at a large volume dedicated center for gastric cancer.

### Patient and Public Involvement

As per local rules and regulations, it was not appropriate or possible to involve patients or the public in the design, or conduct, or reporting, or dissemination plans of our research.

### Inclusion criteria

Sex: Any

Age: 18-80 years old

Histologically confirmed adenocarcinoma

Nonobstructive tumor of the stomach or esophagogastric junction

Clinical stage: cTNM: cT3-4bN1-3M0

Performance status: Eastern Cooperative Oncology Group (ECOG ≤ 2)

Adequate hematological function, liver function, and renal function

Adequate heart and pulmonary function

Written informed consent from the patient

### Exclusion criteria

Acute infectious diseases

Uncontrolled systemic disease or comorbidities

Confirmed or highly suspicious distant metastases

Confirmed or highly suspicious retroperitoneal lymph node metastases

Locally invaded irresectable tumors

Recurrent gastric cancer

Secondary malignant disease

Prior chemo- or radiotherapy

Inclusion in another clinical trial

Known contraindications or hypersensitivity to chemotherapeutic agents

### Reasons for drop out

Protocol violation

Unable to complete planned chemotherapy or surgery for any reason

Refusal to undergo surgery at the same hospital

Consent withdrawn by the participant for any reason

### Ethics

The study was performed according to the Declaration of Helsinki and Good Clinical Practice Guidelines as defined by the International Conference on Harmonisation. The institutional review board approved the study protocol, and patients gave written informed consent for the planned treatment. This study was monitored by the Clinical Research Center (official body responsible for guiding and monitoring all types of research in the hospital), and a timely meeting was performed to check the implementation of protocol guidelines.

### Randomization

Patients were 1:1 randomly assigned to either the FLOT or the SOX group. A blinded statistician at the Clinical Research Center of the hospital was responsible for randomization. The randomization sequence was generated with SPSS software and labeled with random names for different groups. The assignment was made by telephone contact or text messages after the patient met the inclusion criteria and signed the informed consent form.

### Pretreatment assessment

All patients completed all the routine tests, including, but not limited to, the following:

1. Complete blood count, liver and renal function test, clotting analysis, serum tumor biomarkers.
2. Electrocardiography, echocardiography, plain chest radiography.
3. Upper gastrointestinal endoscopy and biopsy for pathological diagnosis.
4. Enhanced computed tomography of the chest, abdomen, and pelvis. CT examination included arterial, venous, and portal phases. CT images consisted of transverse, sagittal, and coronary sections.
5. For suspicious distant metastases, supraclavicular lymph nodes or retroperitoneal lymph nodes on CT, we performed ultrasound tests or magnetic resonance imaging (MRI) as appropriate.
6. Patients underwent bone scintigraphy or positron emission tomography (PET) for suspicious lesions on CT examination.
7. Finally, the diagnosis of peritoneal metastases was made by exploratory laparoscopy.
8. For clinical staging of the disease, we followed the eighth edition of the tumor–node–metastasis (TNM) classification, issued by the International Union against Cancer (UICC).

### Neoadjuvant Chemotherapy

Patients were transferred to the Department of Medical Oncology for chemotherapy, a standard protocol for chemotherapy was circulated, and timely inspection was performed by the investigators and members of the Clinical Research Center to evaluate the implementation of the protocol. Antiemetic drugs with dexamethasone were routinely administered intravenously before chemotherapy. Other supportive drugs, including granulocyte colony-stimulating factor, were given for treatment purposes only. Surgical intervention was allowed for an emergency, e.g., acute upper gastrointestinal bleeding or perforation.

Patients in the FLOT group received four cycles of standard FLOT chemotherapy (7), and the patients in the SOX group received three cycles of

S-1 plus oxaliplatin before curative gastrectomy.

A cycle of FLOT chemotherapy consists of the following:

Day 1: Intravenous 5-fluorouracil (5-FU) 2600 mg/m^2^ via peripherally inserted central catheter (PICC) continued for 24 hours

Intravenous leucovorin 200 mg/m^2^

Intravenous oxaliplatin 85 mg/m^2^

Intravenous docetaxel 50 mg/m^2^

The next chemotherapy cycle was repeated on the 15th day.

The cycle of SOX chemotherapy consisted of the following:

Day 1: Intravenous oxaliplatin 130 mg/m^2^

Day 1-14: Oral tegafur gimeracil oteracil potassium capsule (S-1) 80 mg/m^2^ twice/day

The next chemotherapy was repeated on the 21st day.

### Evaluation of adverse effects

Adverse effects were recorded according to the National Cancer Institute Common Terminology Criteria for Adverse Events (CTCAE 4.0). Drug dose or timing was adjusted for patients with grade three and above adverse effects. Patients with progressive disease were allowed to have alterations in their treatment. Patients were allowed to withdraw from the study for any reason.

### Tumor restaging

Radiologists followed the guidelines of Response Evaluation Criteria in Solid Tumors (RECIST version 1.1) for comparison of radiological response to neoadjuvant chemotherapy (18). Two specialized radiologists independently evaluated the response rate, and the final result was obtained after reviewing both results.

### Surgery

Patients underwent surgical resection between two and four weeks after the completion of neoadjuvant chemotherapy. Exploratory laparoscopy was routinely performed to rule out peritoneal or distant metastases. Only specialist surgeons for gastric cancer were allowed to perform the surgery, and all surgeons were fully aware of the study protocol. Partial or total gastrectomy with D2 lymphadenectomy was performed according to Japanese gastric cancer treatment guidelines (19). Patients with adjacent involved organs underwent combined resection along with gastrectomy. Combined resection was allowed only if R0 resection could be achieved. Distal gastrectomy with Billroth I gastroduodenostomy or Billroth II gastrojejunostomy with Braun anastomosis or Uncut Roux-en-Y gastrojejunostomy or Roux-en-Y gastrojejunostomy was performed for the tumors located at the antrum or lower part of the stomach body. Total gastrectomy with Roux-en-Y esophagojejunostomy was performed for the proximal or large tumors at the body of the stomach. The extent of the surgery was documented to state whether the procedure was curative or noncurative according to the definition stated in Japanese gastric cancer treatment guidelines (19).

### Pathological assessment

Pathologists were blinded to the allocation types of neoadjuvant chemotherapies. After formalin fixation and paraffin embedding, pathologists performed an immunohistochemical examination of all resected specimens. The routine examination included the tumor type; the depth of tumor invasion; the involved lymph nodes; the resection margins; and the invasion of nerves, lymphatics, or blood vessels. Resection or R status was nominated for curative resection (R0) or noncurative resection (R1 and R2). Pathological examinations also included the following: the measurement of the macroscopically identifiable residual tumor and/or scarring indicating the site of the previous tumor bed. Two specialized pathologists followed the Becker criteria for tumor regression grading (TRG) (16). Any conflicting results were settled after re-examination and discussion among both pathologists and investigators.

### Tumor regression grade (TRG) and Becker criteria

“Grade 1a: Complete tumor regression: 0% residual tumor per tumor bed” “Grade 1b: Subtotal tumor regression: <10% residual tumor per tumor bed” “Grade 2: Partial tumor regression: 10-50% residual tumor per tumor bed” “Grade 3: Minimal or no tumor regression: >50% residual tumor per tumor bed”

### Primary endpoints

Total percentage of patients with pathologically complete tumor regression (TRG1a) and subtotal tumor regression (TRG1b) in the primary tumor

### Sample size

This was an exploratory study. The sample size was estimated empirically. The data analysis cut-off time was set at the completion of surgery for the 55^th^ patient, who met the criteria for per-protocol (PP) analysis.

### Statistical analysis

The primary endpoint was analyzed in the intention-to-treat (ITT) population, defined as all the patients who were randomly assigned to a treatment. Postoperative morbidity and mortality were analyzed in the per-protocol (PP) population, which is the number of patients who had surgery after the completion of all planned neoadjuvant chemotherapy. Comparisons of other factors except primary endpoint were post hoc analyses. The statistical analysis was performed with Statistical Package for Social Science (SPSS) version 22.0 for Windows (SPSS, Inc., Chicago, Illinois). Nonparametric methods were used to test the data with an abnormal distribution. The continuous data are described as the median and range. Categorical data are expressed as frequencies and rates. The chi-squared test or Fisher’s exact test was used to compare the differences in rate between the two groups. All p-values presented are two-sided, and a *p*-value less than 0.05 was considered statistically significant.

## Results

Altogether, 74 patients (40 patients in the FLOT group and 34 patients in the SOX group) were enrolled. Nine patients in the FLOT group and ten patients in the SOX group dropped out for different reasons (Fig. 1). Finally, 55 patients completed the planned chemotherapy and underwent surgical resection. All 74 randomly assigned cases were considered the intention-to-treat (ITT) population, and 55 patients who had surgery were considered the per-protocol (PP) population.

**Fig. 1.**
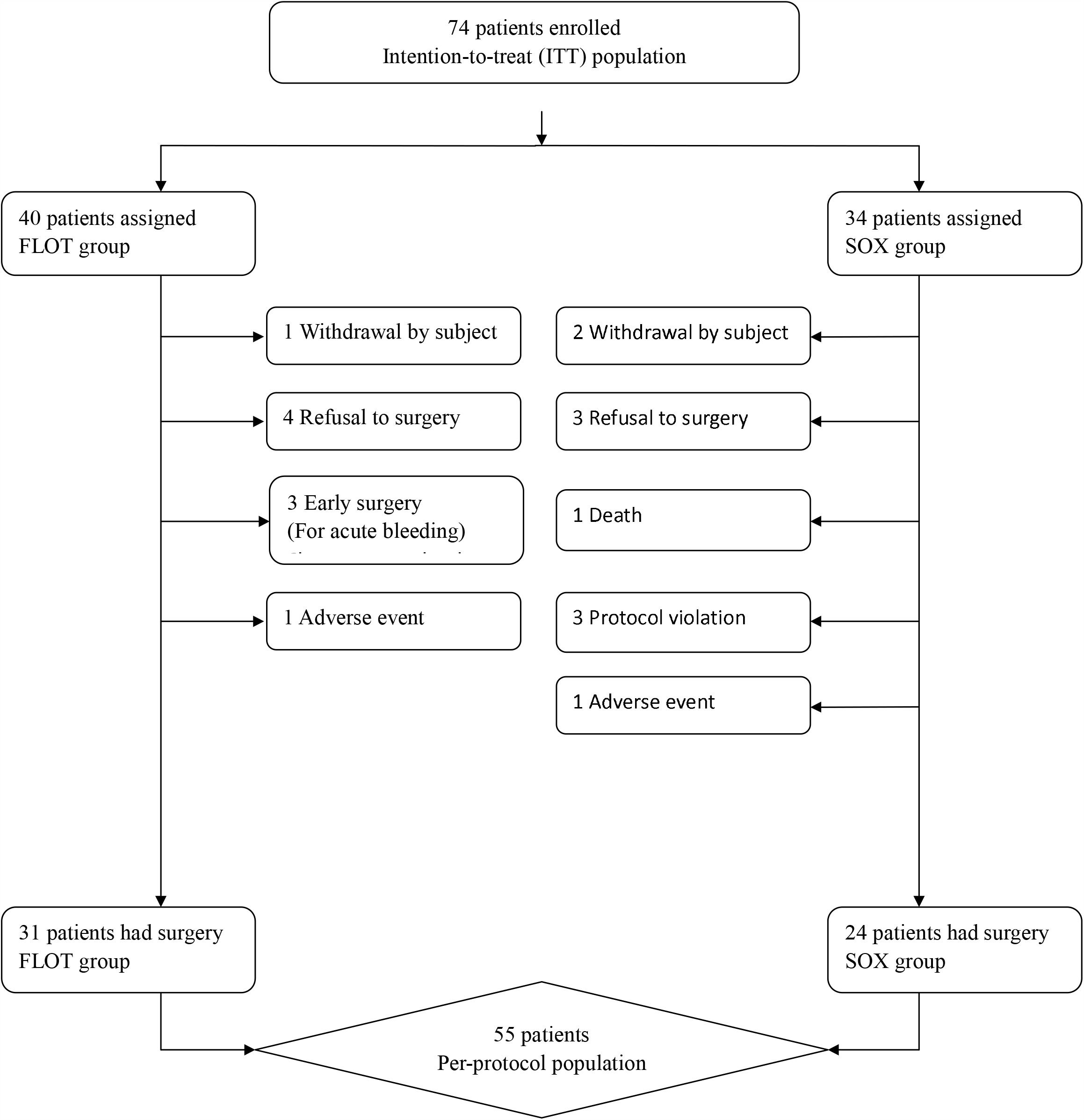
CONSORT diagram.

### Clinical demographics

There was no significant difference in any clinical parameters between the FLOT and SOX groups, including age, sex, or BMI (Table 1, *p*>0.05). There was no significant difference in terms of the location of the tumor or the type of resection. Thirty-two percent of patients in the FLOT group and 33.4% of patients in the SOX group had tumors in the proximal site of the stomach. All patients underwent D2 lymphadenectomy, 58.1% of patients in the FLOT group and 62.5% in the SOX group underwent total gastrectomy (Table 1). One patient in each group underwent pancreatoduodenectomy due to local invasion.

**Table 1.** Demographic data

| Parameter |  | FLOT | SOX | P-Value |
| --- | --- | --- | --- | --- |
| Sex | Male | 29(72.5) | 21(61.8) | 0.326 |
|  | Female | 11(27.5) | 13(38.2) |  |
| Age (Years) | Median | 67 | 61 | 0.155 |
|  | Range | 27-76 | 34-80 |  |
| Body mass index | Median | 23.41 | 23.18 | 0.888 |
|  | Range | 15.69-29.48 | 17.31-27.82 |  |
| Tumor site (PP) | Proximal | 7(22.6) | 7(29.2) | 0.528 |
|  | Proximal-body | 3(9.7) | 1(4.2) |  |
|  | Body | 7(22.6) | 3(12.5) |  |
|  | Distal-body | 6(19.4) | 5(20.8) |  |
|  | Distal | 8(25.8) | 6(25.0) |  |
|  | Total | 0 | 2(8.3) |  |
| Resection type (PP) | Partial | 13(41.9) | 9(37.5) | 0.739 |
|  | Total | 18(58.1) | 15(62.5) |  |

Analysis of intention to treat (ITT) population except otherwise indicated

### Adverse effects

All patients completed 3 cycles of neoadjuvant chemotherapy in the SOX group and 4 cycles of chemotherapy in the FLOT group before surgery. There was no significant difference in chemotherapy-related hematological or nonhematological adverse effects between the two groups (Table 2, *p*>0.05). Most of the hematological or nonhematological adverse events were below grade 3. Nine and five events of hematological grade 3-4 adverse events were observed in the FLOT and SOX groups, respectively (Table 2).

**2.**
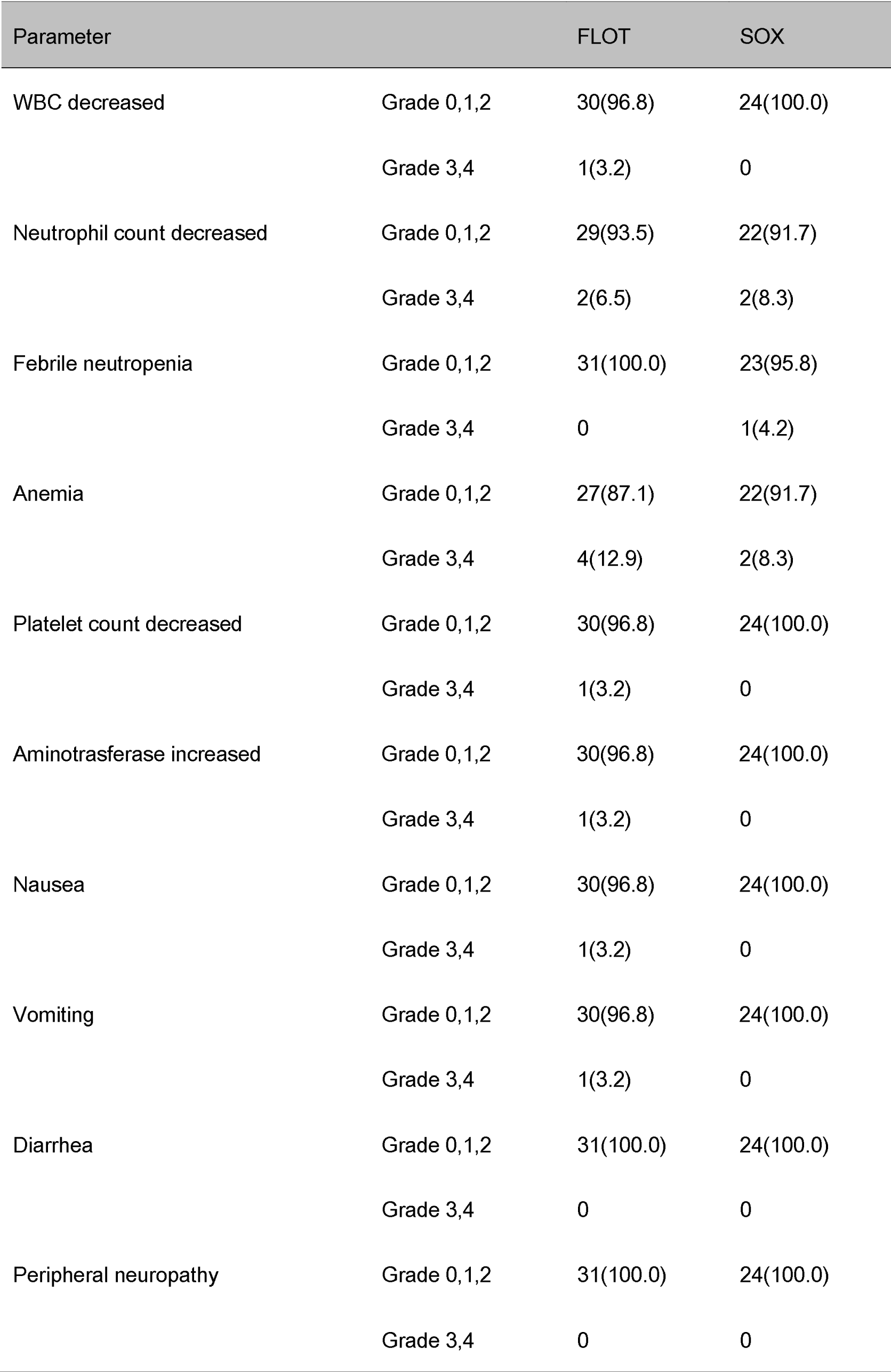
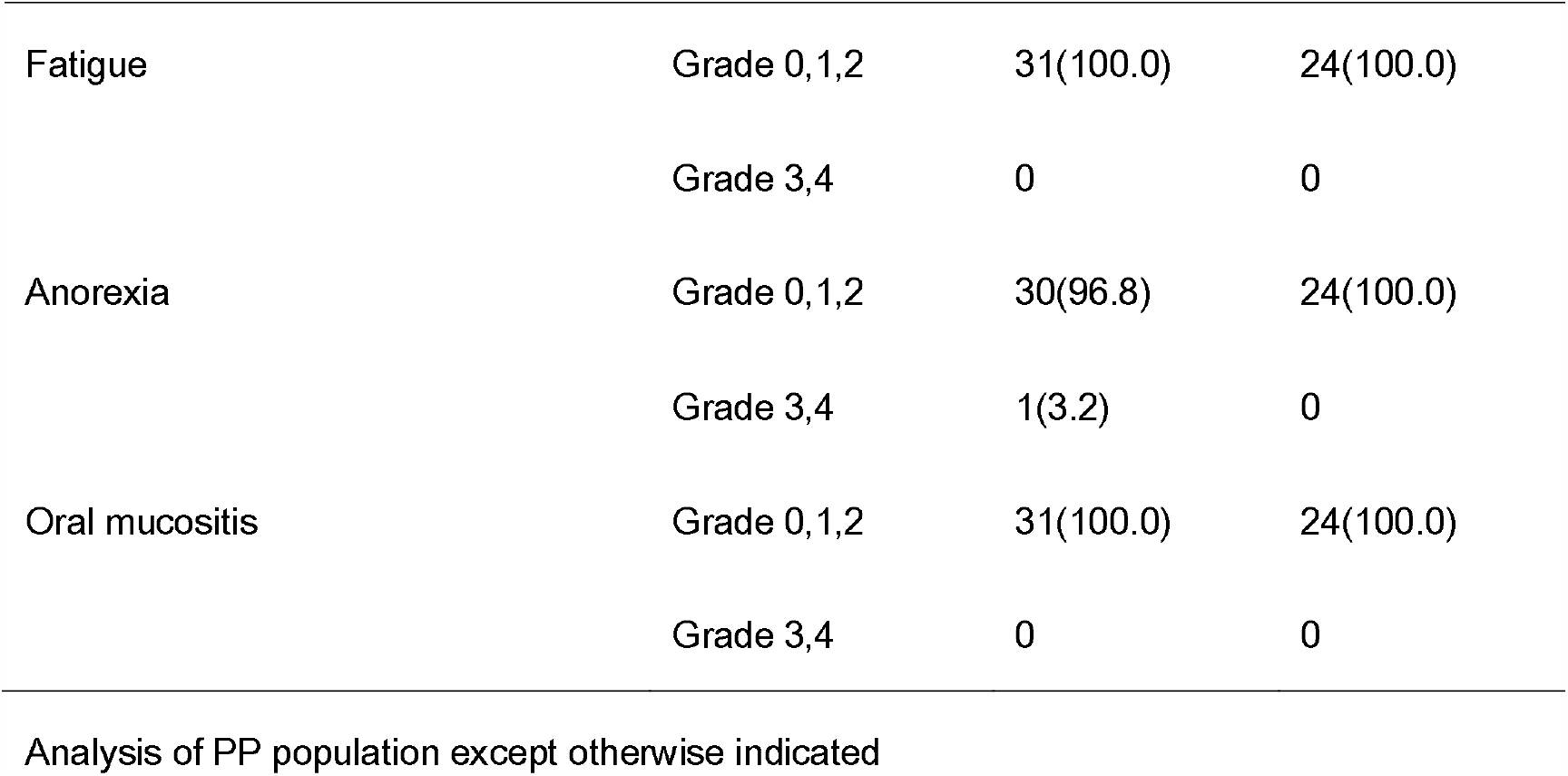
Adverse effects

2. Adverse effects
| Parameter |  | FLOT | SOX |
| --- | --- | --- | --- |
| WBC decreased | Grade 0, 1, 2 | 30(96.8) | 24(100.0) |
|  | Grade 3, 4 | 1(3.2) | 0 |
| Neutrophil count decreased | Grade 0, 1, 2 | 29(93.5) | 22(91.7) |
|  | Grade 3, 4 | 2(6.5) | 2(8.3) |
| Febrile neutropenia | Grade 0, 1, 2 | 31(100.0) | 23(95.8) |
|  | Grade 3, 4 | 0 | 1(4.2) |
| Anemia | Grade 0, 1, 2 | 27(87.1) | 22(91.7) |
|  | Grade 3, 4 | 4(12.9) | 2(8.3) |
| Platelet count decreased | Grade 0, 1, 2 | 30(96.8) | 24(100.0) |
|  | Grade 3, 4 | 1(3.2) | 0 |
| Aminotrasferase increased | Grade 0, 1, 2 | 30(96.8) | 24(100.0) |
|  | Grade 3, 4 | 1(3.2) | 0 |
| Nausea | Grade 0, 1, 2 | 30(96.8) | 24(100.0) |
|  | Grade 3, 4 | 1(3.2) | 0 |
| Vomiting | Grade 0, 1, 2 | 30(96.8) | 24(100.0) |
|  | Grade 3, 4 | 1(3.2) | 0 |
| Diarrhea | Grade 0, 1, 2 | 31(100.0) | 24(100.0) |
|  | Grade 3, 4 | 0 | 0 |
| Peripheral neuropathy | Grade 0, 1, 2 | 31(100.0) | 24(100.0) |
|  | Grade 3, 4 | 0 | 0 |
| Fatigue | Grade 0, 1,2 | 31(100.0) | 24(100.0) |
|  | Grade 3,4 | 0 | 0 |
| Anorexia | Grade 0, 1,2 | 30(96.8) | 24(100.0) |
|  | Grade 3,4 | 1(3.2) | 0 |
| Oral mucositis | Grade 0, 1,2 | 31(100.0) | 24(100.0) |
|  | Grade 3,4 | 0 | 0 |

Analysis of PP population except otherwise indicated

### Radiological response

There was no significant difference in the pretreatment cTNM stage between the FLOT group and the SOX group (Table 3). A total of 41.9% versus 37.5% of cases were diagnosed as stage IVa in the FLOT and SOX groups, respectively. There was no significant difference concerning the radiological response rate between the two groups. In the ITT population, the disease control rate (PR+SD) was comparable between the FLOT group (75.0%) and the SOX group (67.6%), and the overall response rate (ORR) was 55.0% in the FLOT group versus 41.2% in the SOX group (Table 3, *p*>0.05).

**3.**
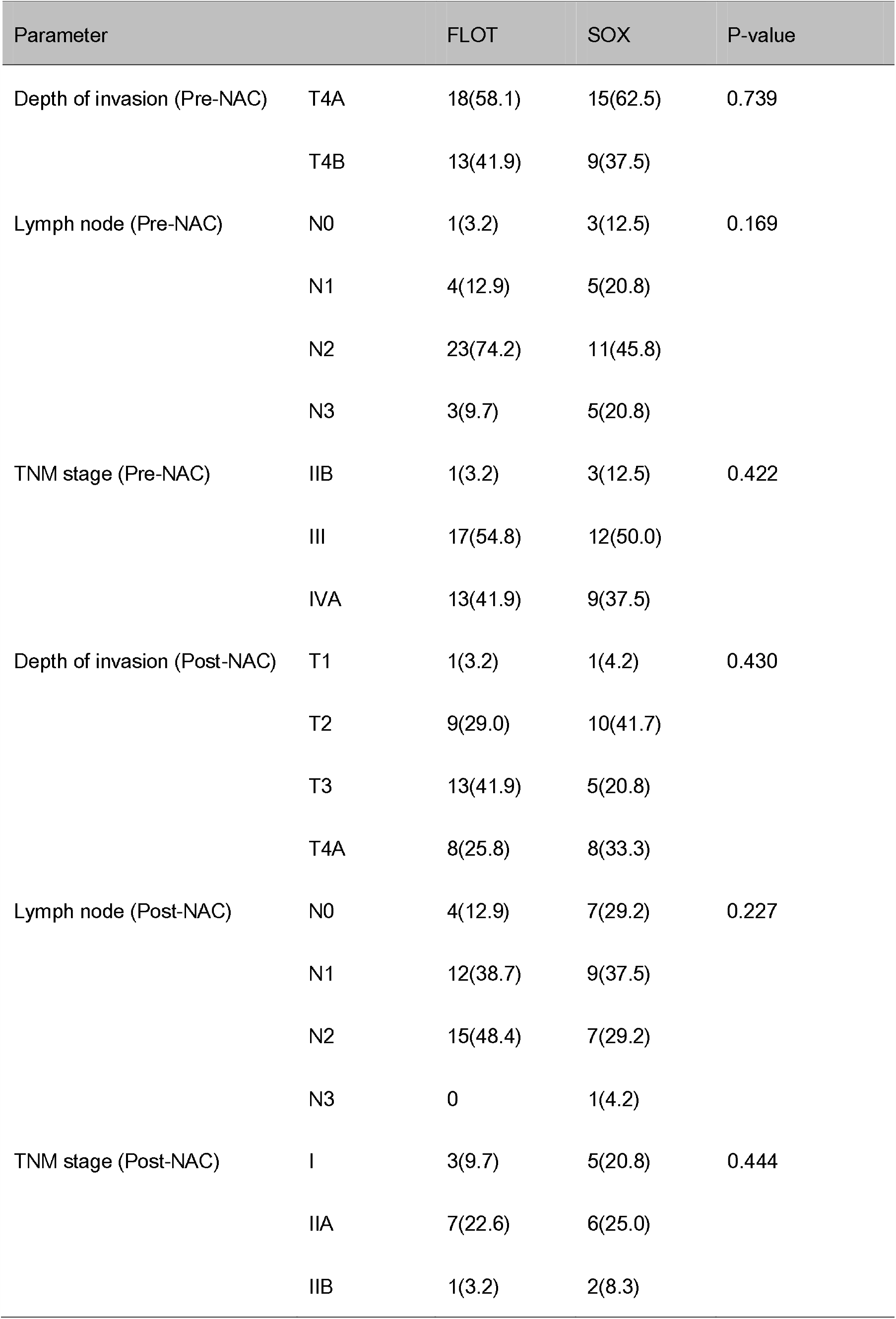
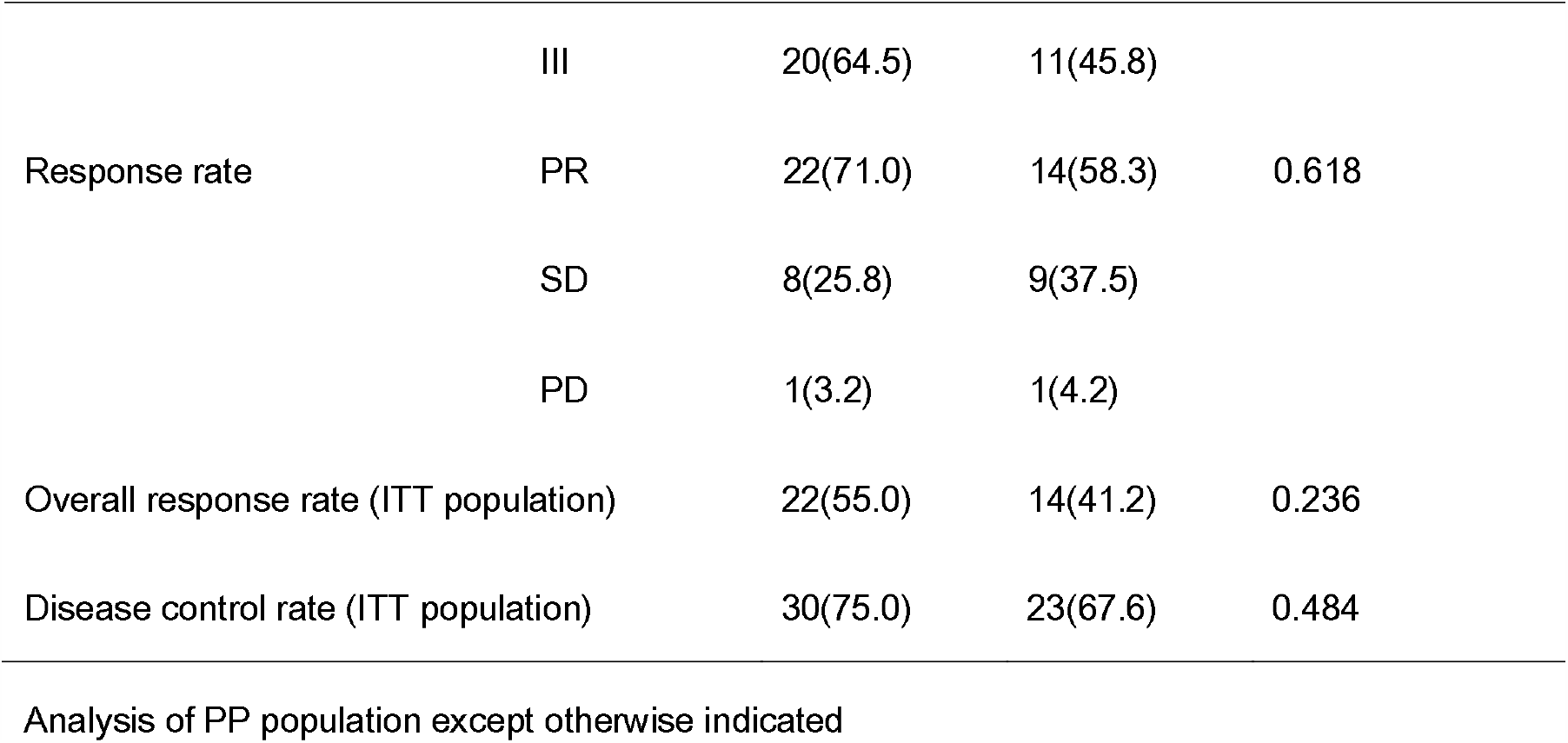
Comparison of CT results between Pre-NAC and Post-NAC

3. Comparison of CT results between Pre-NAC and Post-NAC
| Parameter |  | FLOT | SOX | P-value |
| --- | --- | --- | --- | --- |
| Depth of invasion (Pre-NAC) | T4A | 18(58.1) | 15(62.5) | 0.739 |
|  | T4B | 13(41.9) | 9(37.5) |  |
| Lymph node (Pre-NAC) | N0 | 1(3.2) | 3(12.5) | 0.169 |
|  | N1 | 4(12.9) | 5(20.8) |  |
|  | N2 | 23(74.2) | 11(45.8) |  |
|  | N3 | 3(9.7) | 5(20.8) |  |
| TNM stage (Pre-NAC) | IIB | 1(3.2) | 3(12.5) | 0.422 |
|  | III | 17(54.8) | 12(50.0) |  |
|  | IVA | 13(41.9) | 9(37.5) |  |
| Depth of invasion (Post-NAC) | T1 | 1(3.2) | 1(4.2) | 0.430 |
|  | T2 | 9(29.0) | 10(41.7) |  |
|  | T3 | 13(41.9) | 5(20.8) |  |
|  | T4A | 8(25.8) | 8(33.3) |  |
| Lymph node (Post-NAC) | N0 | 4(12.9) | 7(29.2) | 0.227 |
|  | N1 | 12(38.7) | 9(37.5) |  |
|  | N2 | 15(48.4) | 7(29.2) |  |
|  | N3 | 0 | 1(4.2) |  |
| TNM stage (Post-NAC) | I | 3(9.7) | 5(20.8) | 0.444 |
|  | IIA | 7(22.6) | 6(25.0) |  |
|  | IIB | 1(3.2) | 2(8.3) |  |
|  | III | 20(64.5) | 11(45.8) |  |
| Response rate | PR | 22(71.0) | 14(58.3) | 0.618 |
|  | SD | 8(25.8) | 9(37.5) |  |
|  | PD | 1(3.2) | 1(4.2) |  |
| Overall response rate (ITT population) |  | 22(55.0) | 14(41.2) | 0.236 |
| Disease control rate (ITT population) |  | 30(75.0) | 23(67.6) | 0.484 |

Analysis of PP population except otherwise indicated

### Pathological response

Among the PP population, there was no significant difference in any pathological parameters (Table 4, *p*>0.05). Patients in both groups had favorable margin-free resection: 87.1% in the FLOT group and 100.0% in the SOX group. Lauren’s classification showed that 58.1% of tumors in the FLOT group and 54.2% of tumors in the SOX group were intestinal types. The proportion of T4a tumors and N2 lymph nodes was relatively higher in the FLOT group than in the SOX group, and a greater proportion of postoperative stage III tumors (ypTNM) was observed in the FLOT group than in the SOX group (54.8% versus 45.8%), but there was no significant difference between the two groups. The overall pathological response (TRG grade 1a+1b+2) rate was 67.7% in the FLOT group versus 75% in the SOX group (Table 4, *p*>0.05). In the ITT population, the complete or subtotal TRG was 20% in the FLOT group and 32.4% in the SOX group, but there was no significant difference between the two groups (p>0.05).

**4.**
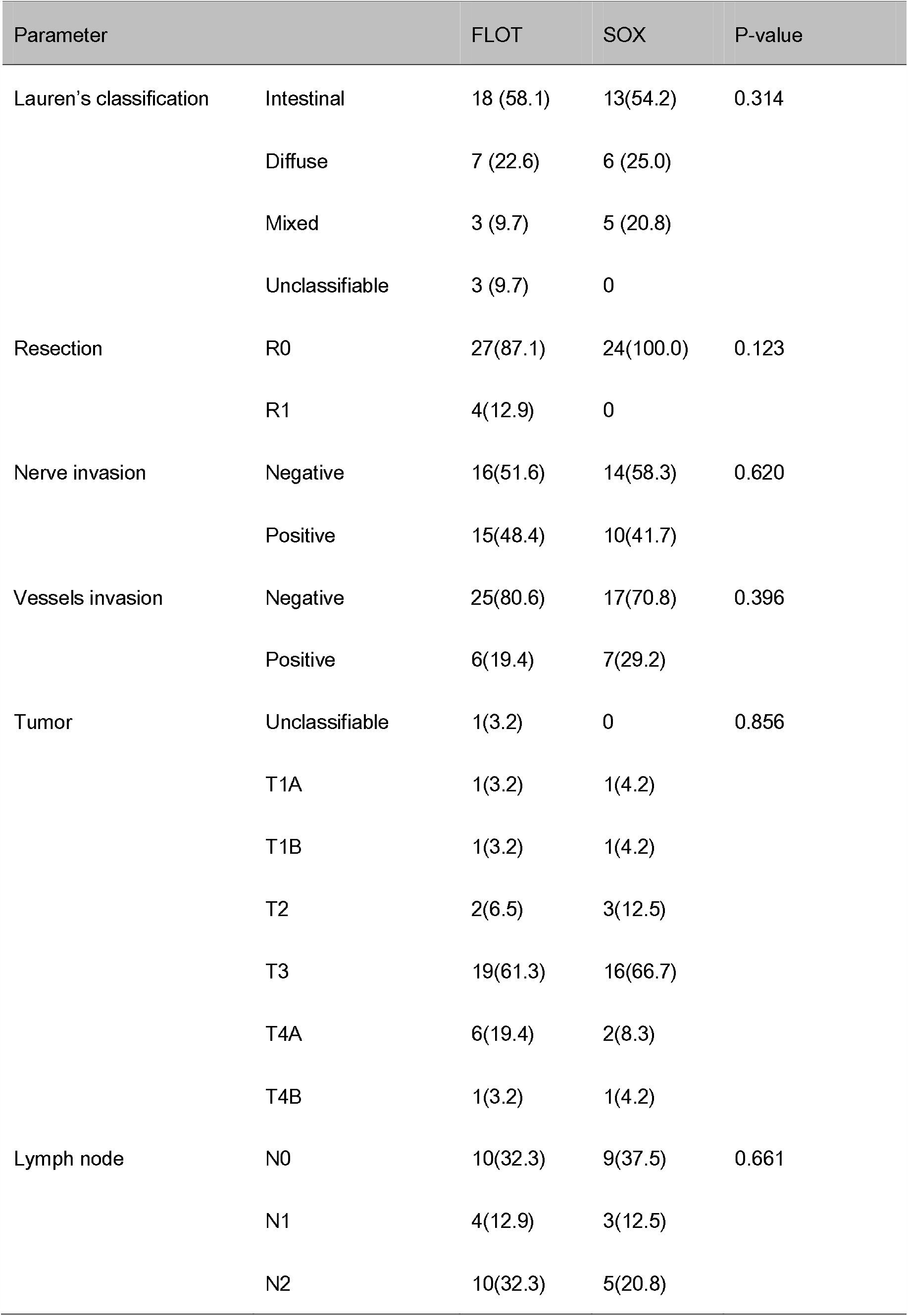
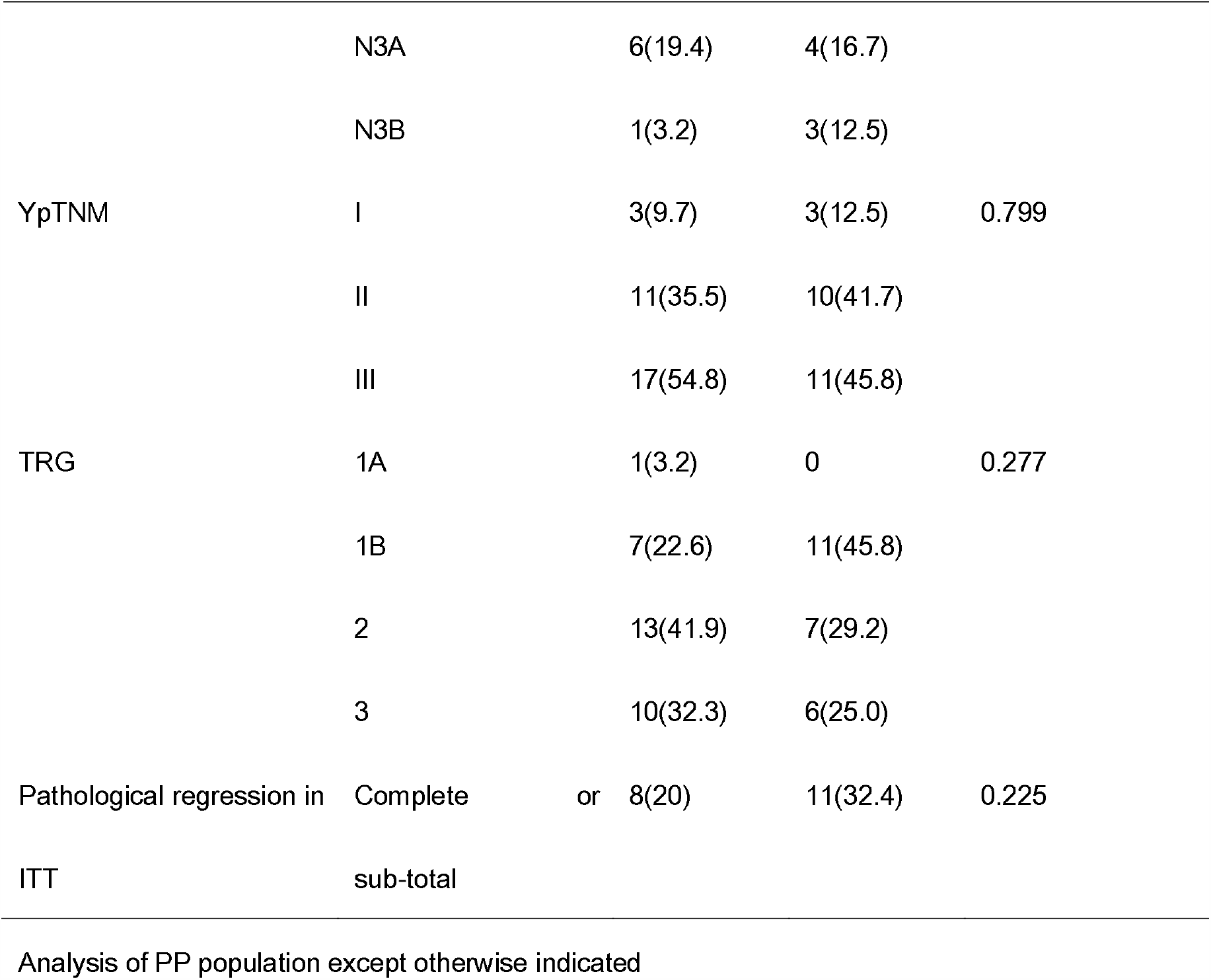
Comparison of postoperative pathological results between two groups

### Postoperative morbidity

There was no significant difference in the postoperative stay at the hospital between the FLOT group and the SOX group, and the median length of stay was 9 days in both groups. There was no significant difference in overall postoperative morbidity between the two groups (Table 5, *p*>0.05). Two (6.5%) anastomotic leakages were observed in the FLOT group and 1 (4.2%) in the SOX group. One patient underwent reoperation for intraabdominal hemorrhage in the SOX group. There were no deaths due to postoperative complications in either group.

**5.** Table Postoperative complications PP population

Analysis of PP population except otherwise indicated 5. Table Postoperative complications PP population
| Complication | Number of patients |  | p value |
| --- | --- | --- | --- |
|  | FLOT | SOX |  |
| Overall complications | 9(29.0) | 8(33.3) | 0.732 |
| Abdominal complication | 7(22.6) | 5(20.8) | 0.876 |
| Intra-abdominal hemorrhage | 0 | 1(4.2) | 0.436 |
| Superficial wound dehiscence | 0 | 1(4.2) | 0.436 |
| Pulmonary Infection | 2(6.5) | 1(4.2) | 1.000 |
| Abdominal Infection | 4(12.9) | 2(8.3) | 0.686 |
| Anastomotic leakage | 2(6.5) | 1(4.2) | 1.000 |
| Pancreatic fistula | 0 | 1(4.2) | 0.436 |
| Impaired renal function | 1(3.2) | 0 | 1.000 |
| Cardiac/respiratory failure | 0 | 1(4.2) | 0.436 |
| Readmission | 1(3.2) | 1(4.2) | 1.000 |
| Reoperation | 0 | 1(4.2) | 0.436 |
| Death | 0 | 0 | NA |

Analysis of PP population except otherwise indicated

## Discussion

Initial reports on the FLOT 4 trial showed that 37% of patients in the FLOT group versus 23% in the ECF/ECX group achieved complete or subtotal tumor regression after neoadjuvant chemotherapy(7). Further results on survival revealed that the patients in the FLOT group had an overall survival of 50 months versus 35 months in patients in the ECF/ECX group (8). Recent small-scale studies from China also suggested that the FLOT regimen was safe and feasible in Chinese patients (9, 10). A propensity-score-matched retrospective study from China also suggested that patients with neoadjuvant FLOT had improved overall survival compared with patients who underwent surgery first(20). The results of these studies suggested that the FLOT regimen was beneficial to locally advanced gastric cancer in terms of pathological regression and survival. However, the combination of fluorouracil and platinum chemoagents, e.g., SOX or XELOX regimens, is commonly used as neoadjuvant chemotherapy in East Asia, including Japan(11-13). Preliminary results of two large-scale RCTs from China (RESOLVE and RESONANCE) further concluded that the SOX regimen is beneficial for LAGC (14, 15). As a result, some controversy remains regarding whether the FLOT regimen is similarly beneficial in East Asian patients. Whether there are any differences between the triplet and the doublet chemoagents in terms of adverse effects and survival benefit has not been studied. In our study, we investigated neoadjuvant FLOT and SOX regimens for patients with LAGC in an attempt to compare the adverse effects and postoperative pathological response between the two groups. Although the sample size is not large enough, to our knowledge, this may be the first head-to-head comparative study of the FLOT and SOX regimens as neoadjuvant chemotherapy for LAGC. The higher proportion of complete or subtotal TRG in the SOX group than in the FLOT group does not indicate the superiority of the SOX regimen over the FLOT regimen because there was no significant difference in terms of statistical calculations. However, at the very least, the results of this study urge the need for further large-scale multicenter randomized trials. Our results may provide some insights for further trials.

There was no difference in the FLOT and SOX groups in terms of adverse effects and postoperative morbidity. Thus, this study further validated the results of our previous study that neoadjuvant chemotherapy with the FLOT regimen is safe in Chinese patients (10). In this study, the demographic data show that patients in both groups were well balanced by randomized assignment. The univariate analysis of all data suggested that TRG was associated with sex, nerve invasion, vessel invasion, and postoperative pathological stage. However, there was no difference in any of these factors between the two groups (Table 4). These data suggest that these known factors, which might have a role in a pathological response, did not influence the results of this study.

There was a conflicting result for the radiological response rate with that of pathological results, which showed that a greater proportion of ORR was seen in the FLOT group than in the SOX group, although there was no statistically significant difference. Therefore, we further analyzed the data according to the stratification of pretreatment clinical staging and postoperative pathological TNM staging. The proportion of complete or subtotal TRG was higher in the SOX group, but there was no significant difference compared to the FLOT group (Table 6).

**Table 6.** Complete or sub-total tumor regression

|  |  | Complete or sub-total tumor regression |  |  |
| --- | --- | --- | --- | --- |
| Stage type | Stage | FLOT | SOX | P VALUE |
| cTNM | IIB | 1/1(100.0) | 1/3(33.3) | NS |
|  | III | 5/17(29.0) | 6/12(50.0) | NS |
|  | IVA | 2/13(15.4) | 4/9(44.4) | NS |
| ypTNM | I | 3/3(100.0) | 3/3(100.0) | NS |
|  | II | 5/11(45.5) | 6/10 (60.0) | NS |
|  | III | 0/17 | 2/11(18.2) | NS |

Analysis of PP population except otherwise indicated
cTNM: Pre-treatment clinical TNM stage;
ypTNM: Postoperative pathological TNM stage

Approximately 37% of patients achieved complete or subtotal TRG in the FLOT4 trial(7), but only 20% of patients achieved complete or subtotal TRG in the FLOT group of our study. Even if we did not compare the result with the SOX group, this result shows that the proportion of complete or subtotal TRG was less than the result of the FLOT4 trial. We hypothesize that the heterogeneity was caused by racial biological differences, and perhaps the FLOT regimen was not as effective in Chinese patients as it was reported to be in German patients. However, concrete data are needed to support this hypothesis. In contrast, the proportion of complete or subtotal TRG in the SOX group was 32.4%, which was comparable to the result of FLOT chemotherapy in the FLOT4 trial (7).

We did not calculate the sample number because there was no good-quality research on tumor regression grading in the SOX regimen, and we had only the results of the FLOT4 trial. Most likely, this was the most important limitation of this study. The number of participants was empirically estimated to obtain preliminary results, and initially, 60 patients were expected to be enrolled for the analysis, but there were a substantial number of patients who dropped out of the trial. Because the primary endpoint of our study was to compare the pathological regression, the cut-off time for data analysis was set at the completion of the surgery of the 55^th^ patient. This cut-off time was set after discussion among investigators and statisticians, without knowing the pathological results. In addition, the results of DFS and OS are still awaited, which will provide further insight into these findings. Of course, further multicenter RCT studies are necessary to elaborate the differences between the FLOT regimen and the SOX regimen as neoadjuvant chemotherapy for patients with LAGC in terms of overall survival.

## Conclusion

Our study demonstrates that the FLOT and SOX regimens are similarly effective for locally advanced gastric cancer patients in terms of clinical downstaging and pathological response. There was no significant difference in adverse effects and postoperative morbidity between the two groups. The results for disease-free survival and overall survival are still awaited. A large-scale phase III multicenter randomized controlled trial is necessary for the validation of this result.

## Data Availability

Any data regarding this study are kept safe with corresponding authors and can be obtained for reasonable request.

## Notes

### Competing Interest Statement

The authors have declared no competing interest.

### Clinical Trial

ClinicalTrials.gov, number NCT03636893

### Funding Statement

The overall costs of publication will be funded by grants from the National Natural Science Foundation of China (No. 91529302 (BY Liu), No.81772509 (Liu BY), and No. 81871904 (Zhu ZG)

### Author Declarations

institutional review board (IRB), Ruijin Hospital

